# Multimodal prediction of 3- and 12-month outcomes in ICU-patients with acute disorders of consciousness

**DOI:** 10.1101/2023.02.06.23285527

**Authors:** Moshgan Amiri, Federico Raimondo, Patrick M. Fisher, Annette Sidaros, Melita Cacic Hribljan, Marwan H. Othman, Ivan Zibrandtsen, Ove Bergdal, Maria Louise Fabritius, Adam Espe Hansen, Christian Hassager, Joan Lilja S Højgaard, Niels Vendelbo Knudsen, Emilie Lund Laursen, Vardan Nersesjan, Miki Nicolic, Karen Lise Welling, Helene Ravnholt Jensen, Sigurdur Thor Sigurdsson, Jacob E. Møller, Jacobo D. Sitt, Christine Sølling, Lisette M. Willumsen, John Hauerberg, Vibeke Andrée Larsen, Martin Ejler Fabricius, Gitte Moos Knudsen, Jesper Kjærgaard, Kirsten Møller, Daniel Kondziella

## Abstract

**Background:** In intensive care unit (ICU) patients with coma and other disorders of consciousness (DoC), outcome prediction is key to decision-making regarding prognostication, neurorehabilitation, and management of family expectations. Current prediction algorithms are largely based on chronic DoC, while multimodal data from acute DoC are scarce. Therefore, CONNECT-ME (Consciousness in neurocritical care cohort study using EEG and fMRI, NCT02644265) investigates ICU-patients with acute DoC due to traumatic and non-traumatic brain injuries, utilizing EEG (resting-state and passive paradigms), fMRI (resting-state) and systematic clinical examinations.

**Methods:** We previously presented results for a subset of patients (*n*=87) concerning prediction of consciousness levels at ICU discharge. Now, we report 3- and 12-month outcomes in an extended cohort (*n*=123). Favourable outcome was defined as modified Rankin Scale ≤3, Cerebral Performance Category ≤2, and Glasgow Outcome Scale-Extended ≥4. EEG-features included visual-grading, automated spectral categorization, and Support Vector Machine (SVM) consciousness classifier. fMRI-features included functional connectivity measures from six resting-state networks. Random-Forest and SVM machine learning were applied to EEG- and fMRI-features to predict outcomes. Here, Random-Forest results are presented as area under the curve (AUC) of receiver operating curves or accuracy. Cox proportional regression with in-hospital death as competing risk was used to assess independent clinical predictors of time to favourable outcome.

**Results:** Between April-2016 and July-2021, we enrolled 123 patients (mean age 51 years, 42% women). Of 82 (66%) ICU-survivors, 3- and 12-month outcomes were available for 79 (96%) and 77 (94%), respectively. EEG-features predicted both 3-month (AUC 0.79[0.77-0.82] and 12-month (0.74[0.71-0.77]) outcomes. fMRI-features appeared to predict 3-month outcome (accuracy 0.69-0.78) both alone and when combined with some EEG-features (accuracies 0.73-0.84), but not 12-month outcome (larger sample sizes needed). Independent clinical predictors of time to favourable outcome were younger age (Hazards-Ratio 1.04[95% CI 1.02-1.06]), TBI (1.94[1.04-3.61]), command-following abilities at admission (2.70[1.40-5.23]), initial brain-imaging without severe pathology (2.42[1.12-5.22]), improving consciousness in the ICU (5.76[2.41-15.51]), and favourable visual-graded EEG (2.47[1.46-4.19]).

**Conclusion:** For the first time, our results indicate that EEG- and fMRI-features and readily available clinical data reliably predict short-term outcome of patients with acute DoC, and EEG also predicts 12-month outcome after ICU discharge.

## Introduction

Since the first report in 2006 of a patient with cognitive-motor dissociation,^1^ the challenge of identifying brain-injured patients with residual consciousness and predicting their long-term recovery has stimulated a new field of research. This, however, mostly concerns patients with subacute or chronic disorders of consciousness (DoC) in rehabilitation facilities.^2,3^

Each year two out of 1000 people fall into a coma and are admitted to an intensive care unit (ICU),^4^ with the key questions being: Who regains consciousness, and who will make a good functional outcome? Accurate prediction of long-term functional outcomes of patients with acute DoC, including coma, is a major challenge, especially during the early phase in the ICU.^5^ While some DoC survivors enter a state of prolonged unresponsive wakefulness, many recover within weeks to months, and a few DoC patients may show signs of recovery even years after the brain injury.^3,6^ Accurate prognostication is hence essential for decision-making in the ICU, including decisions about therapeutic management, withdrawal of life-sustaining therapy,^7–9^ resource allocation and rehabilitation, and management of family expectations. The first step to improve prognostication of acute DoC patients is accurate determination of their levels of consciousness.^10^ This is important since patients with even minimal clinical signs of residual consciousness^11,12^ have more favourable long-term outcomes (as do those with covert consciousness^13–15^). However, determining consciousness levels by routine clinical exams alone is imprecise^16^ because intermittent signs of consciousness are often missed when sensitive systematic ratings scales are omitted.^3,10,17^

In previous work, we established that resting-state EEG, EEG with external stimulations and resting-state fMRI can accurately predict consciousness levels in patients with acute DoC during ICU admission.^18^ Corroborating our findings, multimodal approaches were recommended in a recent review of neuroimaging-based outcome prediction of DoC patients.^19^ However, prognostication of functional recovery of acute DoC is typically limited to unimodal approaches and certain patient subcategories.^6,14,20,21^ Only one study reported 6-month outcome of acute DoC patients with severe traumatic brain injury (TBI) assessed with both EEG and fMRI.^22^ Research reporting the potential of multimodal approaches to predict both early and late functional outcomes of acute DoC patients in the ICU across a wide range of brain injuries is, to our knowledge, non-existent.

To bridge this knowledge gap, we investigated if a multimodal approach consisting of EEG with resting and passive stimulation paradigms, resting-state fMRI, and repeated systematic clinical evaluations could accurately predict functional outcomes of acute DoC patients with TBI and various non-traumatic brain injuries 3 and 12 months after ICU discharge.

## Materials and methods

The CONNECT-ME study (‘Consciousness in neurocritical care cohort study using EEG and fMRI’, NCT02644265) is a prospective, observational, tertiary center cohort, diagnostic phase IIb study. Detailed methods of data acquisition and analysis are described in the study protocol^23^ and a recent paper.^18^ Results concerning the prediction of consciousness levels in a subset of patients (*n=*87) at ICU discharge have been published elsewhere.^18^ Here, we evaluated 3- and 12-month functional outcomes in an extended patient cohort (*n=*123). Below is a brief overview of the methods. **Fig. 1** shows the flow of patients through the study.

**Figure 1.**
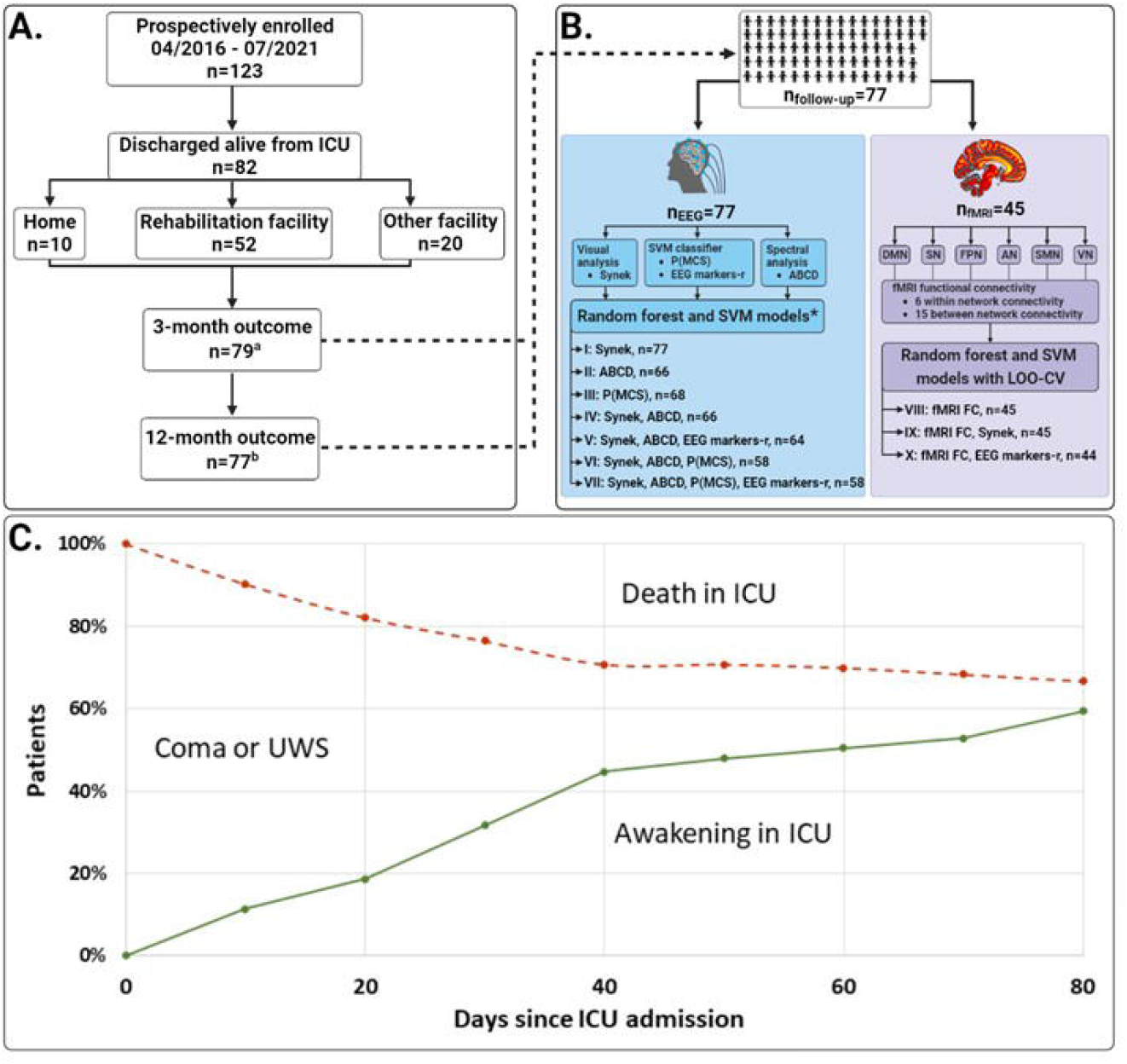
Study flowchart, data assessment strategy and death in ICU. **(A)** 123 acute DoC patients were included, of whom 41 died during ICU admission. Of the 82 patients discharged alive, 10 (12%) patients were discharged directly to their own home, 20 (24%) to other care facilities such as nursing homes, and the remaining 52 (63%) to a high-level neurorehabilitation facility. Three-month follow-up data was available from 79 (96%) patients, and 12-month follow-up data from 77 (94%) patients. **(B)** Full sets of 3- and 12-month follow-up data were available for 77 (94%) patients. EEG recordings were available from all patients (blue box), while fMRI resting-state sequences were available from 45 (58%) patients (purple box). EEGs were analysed with three different approaches; 1) visual manual analysis and scoring according to the Synek scale, 2) automated spectral analysis according to the ABCD model, and 3) a machine learning based SVM consciousness classifier resulting in the probability of being at least in a minimal conscious state (P(MCS)) and 68 EEG markers derived from segments of resting-state EEG (EEG markers-r). Two different machine learning algorithms (i.e., Random-Forest and SVM) were used to conduct seven different predictive models based on EEG features (i.e., models I to VII) and three different models including fMRI features with or without EEG features (i.e., models VIII to X). Models including fMRI features were assessed with additional LOO-CV procedure due to the limited number of available samples. **(C)** This part depicts the proportion of patients in coma or UWS who either awoke to at least MCS-(i.e., regained consciousness to some degree) or died during ICU admission. At time 0 (admission to the ICU) none of the patients were awake (0%) and all were alive (100%). The red line shows the proportion of patients who died in the ICU, and the green line shows the proportion of patients who awoke from coma or UWS in the ICU. During ICU admission, a total of 41 patients (33%) died, while 82 (67%) survived, of whom 73 (59%) awoke prior to ICU discharge. The area between the red and green line indicates the proportion of patients (7%) who remained in coma or UWS at ICU discharge. ^a^ including 8 patients who died prior to 3-month follow-up; ^b^ including 13 patients who died prior to 12-month follow-up; * all EEG models were also tested with same-sample data for head-to-head comparison (see also **Table 3**)

### Patients and study design

We prospectively included patients admitted to one of the four ICUs (excluding the neonatal ICU) at Rigshospitalet (Campus Blegdamsvej), Copenhagen University Hospital, Copenhagen, Denmark between April-2016 and July-2021, and collected demographics, clinical status, and data regarding previous medical history. We included ICU patients with acute DoC (time from brain injury <31 days), age ≥16 years, Danish or English language proficiency, who had a clinical indication for a structural brain MRI ordered by the treating physician. Clinical exams, EEG and fMRI were all performed within a 24-hour window or as close to this time as possible. We aimed for unsedated patients or for the lowest possible sedation levels if patients could not be fully weaned from sedation. Sedation levels were graded as “none or minimal”, “low to moderate”, and “high or very high”.^18^ Patients with contraindications for MRI, major pre-morbid neurological deficits (e.g., mental retardation, aphasia, or deafness), and/or acute life-threatening conditions with immediate risk of clinical deterioration were excluded.

### Classification of consciousness levels

We classified patients according to their level of consciousness into coma^24,25^, unresponsive wakefulness syndrome^26^ (UWS, only reflex behaviour such as spontaneous eye opening), minimally conscious state -/+^27,28^ (MCS-, definite signs of non-reflex behaviour such as visual pursuit, localisation to noxious stimuli or relevant emotional response; MCS+, ability to follow simple commands repeatedly, but not necessarily consistently), emergence from MCS^29^ (eMCS, reliable two-way communication or functional object use) or locked-in syndrome^30^ (LIS; consistent and reliable communication by rudimentary eye opening), applying previously described clinical examination techniques.^18^ Briefly, consciousness levels were determined at the time of enrolment and at ICU discharge using a systematic clinical approach including sub-elements of the coma recovery scale revised (CRS-R)^17^ with the addition of Glasgow Coma Scale (GCS)^31^, and the Full Outline of UnResponsiveness (FOUR).^32^ Furthermore, daily routine neurological exams were performed by the attending team of physicians and results accessed from the electronic health records.

## EEG

Standard 19/25 channel bedside video-EEG (NicoletOne, Natus Medical Inc., Middleton, WI, USA) was recorded with electrodes placed according to the international 10/20 system.^33^ All EEGs contained a 10-minute resting-state segment, and for reactivity assessment a segment with stimulations including eye opening, calling the patient by their name, noxious stimuli applied as pressure to earlobes, fingertips and sternum, and sensory tactile stimuli applied with a cotton swap to the nostrils. Investigators performing EEG analyses were unaware of patient outcomes.

EEGs were assessed in three different ways, as described previously;^18^ (1) manual visual analysis by two experienced board-certified neurophysiologists (MHC and AS) scoring the EEGs according to the Synek scale^34^ (level I to V with increasing level indicating increasing pathology), (2) ABCD spectral analysis as described by Forgacs et al.^35^ (with category “A” indicating complete corticothalamic disruption and “D” full recovery of corticothalamic circuit), and (3) a Support Vector Machine (SVM) based consciousness classifier^36^ predicting the probability of the patient’s consciousness level being at least MCS- (P(MCS)) from 68 EEG markers derived separately from EEG resting segments and segments with stimulations.

### fMRI

A 10-minute resting-state scan session with a T2*-weighted echo-planar imaging BOLD fMRI sequence was performed on 1.5 or 3 Tesla MRI-scanners (Siemens, Erlangen, Germany) with 20-or 64-channel head coils, respectively. Pre-processing of fMRI data was performed using SPM12 in MATLAB v2019a (https://www.fil.ion.ucl.ac.uk/spm/software/spm12/) according to our previously described method.^18^ Briefly, denoised regional timeseries were extracted and region-to-region functional connectivity estimated by calculating the timewise correlation coefficient (Pearson’s rho) between each pair of regional timeseries and applying Fisher’s r-to-z transformation to the correlation coefficient. A total of 21 within- and between-network functional connectivity measures were calculated as the average functional connectivity across the set of respective region-to-region pairs for six resting-state networks (i.e., the default mode network (DMN), frontoparietal network (FPN), auditory network (AN), salience network (SN), sensorimotor network (SMN) and visual network (VN)). Investigators assessing fMRI data were unaware of patient outcomes.

### Follow-up data

We used three outcome scales to assess functional outcome at 3 and 12 months after ICU discharge; 1) modified Rankin Scale (mRS),^37^ 2) Glasgow Outcome Scale Extended (GOS-E)^38^ and 3) Cerebral Performance Category (CPC),^39^ respectively (**Box S1**). The mRS is used for evaluation of recovery in stroke patients with focus on the patient’s ability to walk with or without assistance.^37^ The GOS-E is an overall functional outcome scale frequently used to collect follow-up data of TBI patients and include other aspects of functional recovery such as the ability to work, socialize, and level of emotional deficits.^38^ Finally, the CPC is an evaluation tool to assess level of recovery of cardiac arrest patients with regaining of consciousness considered a main aspect.^39^ By including all three scales, we aimed at evaluating different aspects of functional recovery since our study cohort consists of a heterogeneous group of patients regarding the cause of brain injury (i.e. stroke, TBI, cardiac arrest, and other neurological and medical causes). Functional outcome was determined from electronic health records typically based on structural assessments by experienced nursing staff at the high-level rehabilitation facility most surviving patients were discharged to. If sufficient data was not available from health records; patients, family members or other caregivers were contacted by telephone. Favourable outcome was defined as a combination of mRS ≤3 (indicating that patients can walk unassisted), GOS-E ≥4 (indicating that patients can take care of themselves alone for at least 8 hours at home) and CPC ≤2 (indicating that patients are conscious and independent of others for activities of daily living). Patients who died after hospital discharge were included in primary outcome analysis, while patients who died during ICU were excluded as were patients lost to follow-up.

### Machine learning algorithms and predictive models

Two machine learning algorithms where utilized: Random-Forest and Support Vector Machine (SVM). This ensured exploiting both linear and non-linear interactions. Algorithms were trained to predict binary outcome at 3- and 12-month follow-up. Model’s performance was estimated using stratified 5-fold cross-validation (repeated 10 times). A special cross-validation scheme (leave-one-out cross-validation,^40^ LOO-CV) was used to evaluate the potential of fMRI-features, since the limited fMRI samples available from patients with follow-up outcome did not allow to obtain reliable estimates with 5-fold CV. Algorithms’ hyperparameters were selected using nested-cross validation and a grid-search procedure. Both unimodal models based on single features (EEG- or fMRI-features) and multimodal models based on a combination of several features (e.g., combination of EEG- and fMRI-features) were developed with main outcome measures as binary targets. In total, ten different predictive models (I-X) were developed and tested with each algorithm. Same-sample models were tested for head-to-head comparison of EEG-features but could not be tested with fMRI-features due to the low number of available patients with the full set of EEG-features, fMRI-features, and outcome measures. Prediction performance of models evaluated with 5-fold CV were assessed with area under the curve (AUC) of receiver operating characteristic (ROC) curves, sensitivity and positive predictive value (PPV), while performance of the LOO-CV models including fMRI-features were assessed with the accuracy measure (ratio of correctly predicted samples over total samples). AUC, sensitivity and PPV estimates are reported as mean [95% CI] and accuracies as a number between 0-1. The models hence predict the precision with which favourable outcomes can be distinguished from unfavourable outcomes. All machine learning analyses were done using Julearn and scikit-learn.^41^

### Outcome measures

Our primary target outcome was binary outcome at 3- and 12-month follow-up. Time to favourable outcome was considered a secondary outcome.

### Statistical analysis

Quantitative data are expressed as mean ± standard deviation (SD) or median (IQR) and group comparisons assessed with Student t-test, Mann-Whitney-U, or Kruskal-Wallis test. Categorical data are expressed as numbers (percentages) and compared using chi-squared test or Fisher’s exact test. Cox proportional hazards regression model with in-hospital death considered as a competing risk is used for the assessment of important predictors of time to favourable outcome. Multicollinearity analysis was performed, and variable inflation factor assessed to avoid high level of correlation between the variables in the regression model. Results are expressed as hazards ratio (HR) with corresponding 95% confidence intervals (CI) and p-values. The statistical software R version 4.2.0 (2022-04-22) was used for statistical analysis.

### Data availability

fMRI data cannot be made fully anonymous and are not publicly available. Other data will be shared upon reasonable request. The code used in the predictive models is available at https://github.com/fraimondo/connectme-followup.

### Ethics

This study was approved by The Danish Data Protection Agency (RH-2016-191, I-Suite nr:04760) and the Ethics Committee of the Capital Region of Denmark (File-nr.:H-16040845). Written consent was waived because all data were acquired during routine clinical work-up. CONNECT-ME is registered with clinicaltrials.org (NCT02644265).

## Results

### Demographics and clinical characteristics

We included 123 patients (mean age 51±19 years; 51 (42%) women), of whom 82 (67%) were discharged alive from the ICU (**Fig. 1 and Table 1**). Of the 41 deaths in the ICU, 37 (90%) occurred after withdrawal of life-sustaining therapy. Advanced age, preadmission comorbidity, cardiac arrest as cause of ICU admission (OR 10.4 [2.46-78.3]), lower GCS motor score at admission, lower total GCS and FOUR score at study enrolment, lower consciousness levels at study enrolment and shorter duration of ICU admission were all significantly associated with death in the ICU (all *P* < 0.05, see also **Table 1**). EEG was available from 122 (99%) patients, while fMRI was available from 67 (54%) patients. The proportion of patients with fMRI did not differ between those who died in the ICU and patients discharged alive.

**Table 1.**
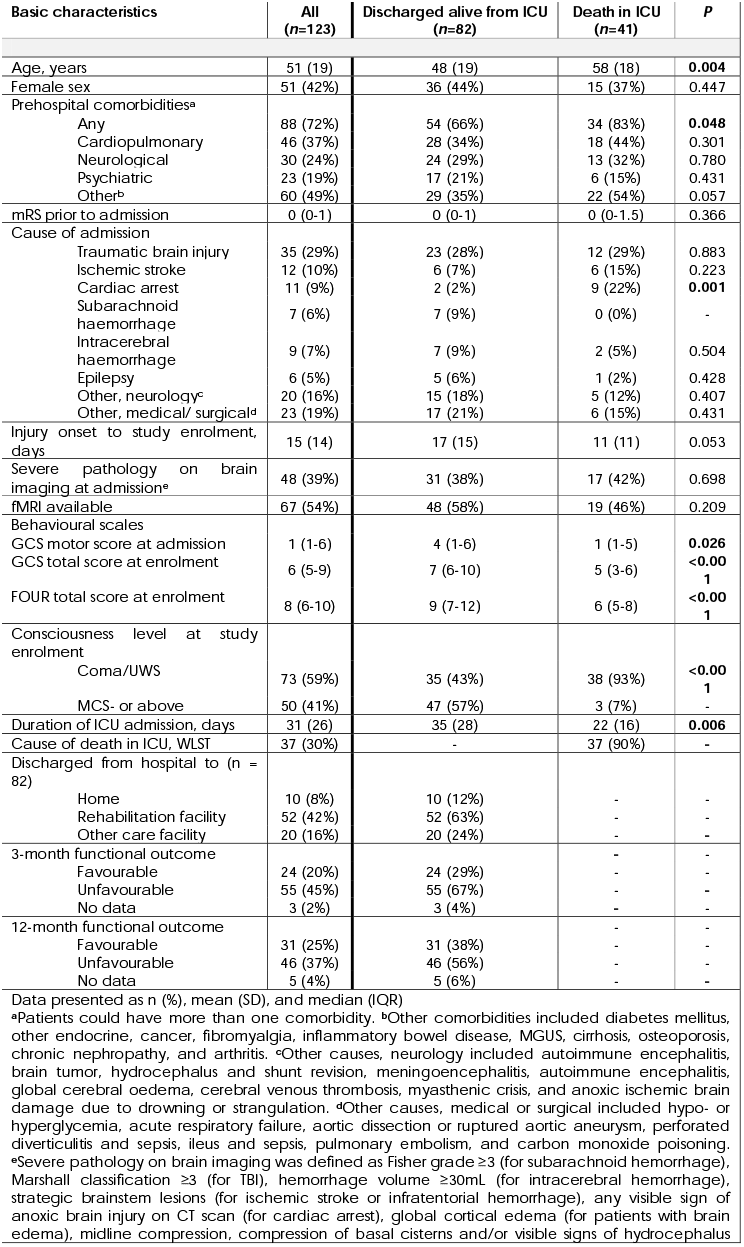
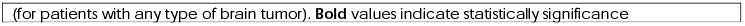
Demographics and clinical characteristics of study population, and comparison of patients discharged alive from patients who died in the ICU.

| Basic characteristics | All<br>(n=123) | Discharged alive from ICU<br>(n=82) | Death in ICU<br>(n=41) | P |
| --- | --- | --- | --- | --- |
| Age, years | 51 (19) | 48 (19) | 58 (18) | **0.004** |
| Female sex | 51 (42%) | 36 (44%) | 15 (37%) | 0.447 |
| Prehospital comorbidities<sup>a</sup> |  |  |  |  |
| Any | 88 (72%) | 54 (66%) | 34 (83%) | **0.048** |
| Cardiopulmonary | 46 (37%) | 28 (34%) | 18 (44%) | 0.301 |
| Neurological | 30 (24%) | 24 (29%) | 13 (32%) | 0.780 |
| Psychiatric | 23 (19%) | 17 (21%) | 6 (15%) | 0.431 |
| Other<sup>b</sup> | 60 (49%) | 29 (35%) | 22 (54%) | 0.057 |
| mRS prior to admission | 0 (0-1) | 0 (0-1) | 0 (0-1.5) | 0.366 |
| Cause of admission |  |  |  |  |
| Traumatic brain injury | 35 (29%) | 23 (28%) | 12 (29%) | 0.883 |
| Ischemic stroke | 12 (10%) | 6 (7%) | 6 (15%) | 0.223 |
| Cardiac arrest | 11 (9%) | 2 (2%) | 9 (22%) | **0.001** |
| Subarachnoid haemorrhage | 7 (6%) | 7 (9%) | 0 (0%) | - |
| Intracerebral haemorrhage | 9 (7%) | 7 (9%) | 2 (5%) | 0.504 |
| Epilepsy | 6 (5%) | 5 (6%) | 1 (2%) | 0.428 |
| Other, neurology<sup>c</sup> | 20 (16%) | 15 (18%) | 5 (12%) | 0.407 |
| Other, medical/ surgical<sup>d</sup> | 23 (19%) | 17 (21%) | 6 (15%) | 0.431 |
| Injury onset to study enrolment, days | 15 (14) | 17 (15) | 11 (11) | 0.053 |
| Severe pathology on brain imaging at admission<sup>e</sup> | 48 (39%) | 31 (38%) | 17 (42%) | 0.698 |
| fMRI available | 67 (54%) | 48 (58%) | 19 (46%) | 0.209 |
| Behavioural scales |  |  |  |  |
| GCS motor score at admission | 1 (1-6) | 4 (1-6) | 1 (1-5) | **0.026** |
| GCS total score at enrolment | 6 (5-9) | 7 (6-10) | 5 (3-6) | **<0.001** |
| FOUR total score at enrolment | 8 (6-10) | 9 (7-12) | 6 (5-8) | **<0.001** |
| Consciousness level at study enrolment |  |  |  |  |
| Coma/UWS | 73 (59%) | 35 (43%) | 38 (93%) | **<0.001** |
| MCS- or above | 50 (41%) | 47 (57%) | 3 (7%) | - |
| Duration of ICU admission, days | 31 (26) | 35 (28) | 22 (16) | **0.006** |
| Cause of death in ICU, WLST | 37 (30%) | - | 37 (90%) | - |
| Discharged from hospital to (n = 82) |  |  |  |  |
| Home | 10 (8%) | 10 (12%) | - | - |
| Rehabilitation facility | 52 (42%) | 52 (63%) | - | - |
| Other care facility | 20 (16%) | 20 (24%) | - | - |
| 3-month functional outcome |  |  |  |  |
| Favourable | 24 (20%) | 24 (29%) | - | - |
| Unfavourable | 55 (45%) | 55 (67%) | - | - |
| No data | 3 (2%) | 3 (4%) | - | - |
| 12-month functional outcome |  |  |  |  |
| Favourable | 31 (25%) | 31 (38%) | - | - |
| Unfavourable | 46 (37%) | 46 (56%) | - | - |
| No data | 5 (4%) | 5 (6%) | - | - |

Data presented as n (%), mean (SD), and median (IQR)
<sup>a</sup>Patients could have more than one comorbidity. <sup>b</sup>Other comorbidities included diabetes mellitus, other endocrine, cancer, fibromyalgia, inflammatory bowel disease, MGUS, cirrhosis, osteoporosis, chronic nephropathy, and arthritis. <sup>c</sup>Other causes, neurology included autoimmune encephalitis, brain tumor, hydrocephalus and shunt revision, meningoencephalitis, autoimmune encephalitis, global cerebral oedema, cerebral venous thrombosis, myasthenic crisis, and anoxic ischemic brain damage due to drowning or strangulation. <sup>d</sup>Other causes, medical or surgical included hypo- or hyperglycemia, acute respiratory failure, aortic dissection or ruptured aortic aneurysm, perforated diverticulitis and sepsis, ileus and sepsis, pulmonary embolism, and carbon monoxide poisoning. <sup>e</sup>Severe pathology on brain imaging was defined as Fisher grade $\geq 3$ (for subarachnoid hemorrhage), Marshall classification $\geq 3$ (for TBI), hemorrhage volume $\geq 30$ mL (for intracerebral hemorrhage), strategic brainstem lesions (for ischemic stroke or infratentorial hemorrhage), any visible sign of anoxic brain injury on CT scan (for cardiac arrest), global cortical edema (for patients with brain edema), midline compression, compression of basal cisterns and/or visible signs of hydrocephalus
(for patients with any type of brain tumor). **Bold** values indicate statistical significance

### Functional outcome and time to favourable outcome

Of the 82 patients discharged alive from the ICU, functional outcome was available from 79 (96%) at 3 months and from 77 (94%) at 12 months (**Fig. 1**). Thirteen patients (16%) died prior to 12-month follow-up, of whom eight were dead by 3-month follow-up. Of the 79 patients with 3-month follow-up data, 26 (33%) had a mRS score of ≤3, 24 (30%) a CPC score of ≤2 and 33 (42%) a GOS-E score of ≥4. Of the 77 patients with 12-month follow-up data, 32 (42%) had a mRS score ≤3, 33 (43%) a CPC score ≤2, and 44 (57%) a GOS-E score ≥4. Overall, 24 (30%) of the 79 patients had favourable outcome (i.e., favourable functional outcome according to all three outcome scales) at 3 months, and 31 (40%) of 77 at 12 months. Patients with an unfavourable outcome at both 3 and 12 months were more likely to be discharged from hospital to a high-level rehabilitation facility or another care facility such as a nursing home rather than to their own home. Clinical characteristics and comparison of patients with favourable and unfavourable 3- and 12-month functional outcomes are shown in **Table 2**. As illustrated by **Fig. 2**, variables independently predicting time to favourable outcome were younger age (HR 1.04 [95% CI 1.02-1.06]), TBI as cause of ICU admission (HR 1.94 [1.04-3.61]), ability to follow commands at admission (HR 2.70 [1.40-5.23]), improving consciousness level during the stay in the ICU (HR 5.76 [2.14-15.51]) and initial brain-imaging without severe pathology (HR 2.42 [1.12-5.22]). Furthermore, favourable visual EEG grading (i.e., Synek score I or II) (HR 2.47 [1.46-4.19]) was also an independent predictor of time to favourable outcome (**Fig. 2**).

**Table 2.** Comparison of patients with favourable and unfavourable 3- and 12-month functional outcomes.

|  |  |  |  |  |  |  |
| --- | --- | --- | --- | --- | --- | --- |
| Basic characteristics |  |  |  |  |  |  |
|  | 3-month outcome (n=79) |  |  | 12-month outcome (n=77) |  |  |
|  | Favourable (n=24) | Unfavourable (n=55) | P | Favourable (n=31) | Unfavourable (n=46) | P |
| Age, years | 43 (19) | 49 (19) | 0.189 | 38 (16) | 54 (18) | <0.001 |
| Female sex | 11 (46%) | 24 (44%) | 0.858 | 14 (45%) | 21 (46%) | 0.968 |
| Preadmission comorbidities<sup>a</sup> |  |  |  |  |  |  |
| Any | 16 (67%) | 36 (66%) | 0.927 | 17 (55%) | 35 (76%) | 0.059 |
| Cardiopulmonary | 7 (29%) | 19 (35%) | 0.658 | 6 (19%) | 20 (44%) | **0.030** |
| Neurological | 3 (13%) | 21 (38%) | **0.022** | 5 (16%) | 19 (42%) | **0.021** |
| Psychiatric | 10 (42%) | 7 (13%) | **0.007** | 11 (36%) | 6 (13%) | **0.026** |
| Other medical<sup>b</sup> | 8 (33%) | 20 (36%) | 0.809 | 7 (23%) | 21 (46%) | **0.042** |
| mRS prior to admission | 0 (0-0.5) | 0 (0-1) | 0.098 | 0 (0-0) | 1 (0-2) | **0.005** |
| Clinical characteristics |  |  |  |  |  |  |
| Cause of admission |  |  |  |  |  |  |
| Traumatic brain injury | 6 (25%) | 16 (29%) | 0.729 | 10 (32%) | 11 (24%) | 0.434 |
| Ischemic stroke | 1 (4%) | 5 (9%) | 0.509 | 2 (6%) | 4 (9%) | 0.758 |
| Cardiac arrest | 0 (0%) | 2 (4%) | 1.000 | 0 (0%) | 2 (4%) | 0.513 |
| Subarachnoid haemorrhage | 0 (0%) | 7 (13%) | 0.094 | 1 (3%) | 6 (13%) | 0.165 |
| Intracerebral haemorrhage | 0 (0%) | 7 (13%) | 0.094 | 0 (0%) | 7 (15%) | **0.037** |
| Epilepsy | 2 (8%) | 3 (5%) | 0.644 | 1 (3%) | 4 (9%) | 0.395 |
| Other, neurology<sup>c</sup> | 8 (33%) | 7 (13%) | **0.045** | 8 (26%) | 7 (15%) | 0.270 |
| Other, medical/ surgical<sup>d</sup> | 7 (29%) | 8 (15%) | 0.150 | 9 (29%) | 5 (11%) | 0.054 |
| Severe pathology on brain imaging at admission<sup>e</sup> | 3 (13%) | 28 (51%) | **0.001** | 6 (19%) | 25 (54%) | **0.002** |
| fMRI available | 11 (46%) | 35 (64%) | 0.153 | 19 (61%) | 27 (59%) | 0.827 |
| Behavioural scales |  |  |  |  |  |  |
| GCS motor score at admission | 6 (4-6) | 3 (1-6) | **0.013** | 5 (1-6) | 4 (1-6) | 0.500 |
| GCS total score at enrolment | 6.5 (5-10) | 7 (6-10) | 0.492 | 8 (5-10) | 7 (6-9.75) | 0.762 |
| FOUR total score at enrolment | 8.5 (7-12) | 9 (7-12) | 0.306 | 9 (7-12) | 8.5 (7-12) | 0.243 |
| Consciousness level at study enrolment |  |  | 0.878 |  |  | 0.388 |
| Coma/UWS | 10 (42%) | 24 (44%) | - | 11 (36%) | 21 (46%) | - |
| MCS- or above | 14 (58%) | 31 (56%) | - | 20 (65%) | 25 (54%) | - |
| Consciousness level at ICU discharge |  |  | 0.104 |  |  | **0.023** |
| Coma/UWS | 1 (4%) | 10 (18%) | - | 1 (3%) | 10 (22%) | - |
| MCS- or above | 23 (96%) | 45 (82%) | - | 30 (97%) | 36 (78%) | - |
| Duration of ICU admission, days | 24 (20) | 41 (31) | **0.022** | 31 (30) | 39 (28) | 1.01 (0.99-1.03) |
| Discharged from hospital to |  |  | <0.001 |  |  | **0.009** |
| Home | 9 (38%) | 1 (2%) | - | 8 (26%) | 1 (2%) | - |
| Rehabilitation facility | 6 (25%) | 10 (19%) | - | 5 (16%) | 11 (24%) | - |
| Other care facility | 9 (38%) | 43 (80%) | - | 18 (58%) | 33 (73%) | - |
| Data presented as n (%), mean (SD), and median (IQR) |  |  |  |  |  |  |
| <sup>a</sup>Patients could have more than one comorbidity. <sup>b</sup>Other comorbidities included diabetes mellitus, other endocrine, cancer, fibromyalgia, inflammatory bowel disease, MGUS, cirrhosis, osteoporosis, chronic nephropathy and arthritis. <sup>c</sup>Other causes, neurology included autoimmune encephalitis, brain tumor, hydrocephalus and shunt revision, meningoencephalitis, autoimmune encephalitis, global cerebral oedema, cerebral venous thrombosis, myasthenic crisis, and anoxic ischemic brain damage due to drowning or strangulation. <sup>d</sup>Other causes, medical or surgical included hypo- or hyperglycemia, acute respiratory failure, aortic dissection or ruptured aortic aneurysm, perforated diverticulitis and sepsis, ileus and sepsis, pulmonary embolism, and carbon monoxide poisoning. <sup>e</sup>Severe pathology on brain imaging was defined as Fisher grade ≥3 (for subarachnoid hemorrhage), Marshall classification ≥3 (for traumatic brain injury), hemorrhage volume ≥30mL (for intracerebral hemorrhage), strategic brainstem lesions (for ischemic stroke or intratentorial hemorrhage), any visible sign of anoxic brain injury on CT scan (for cardiac arrest), global cortical edema (for patients with brain edema), midline compression, compression of basal cisterns and/or visible signs of hydrocephalus (for patients with any type of brain tumor). **Bold** values indicate statistically significance |  |  |  |  |  |  |

**Figure 2.**
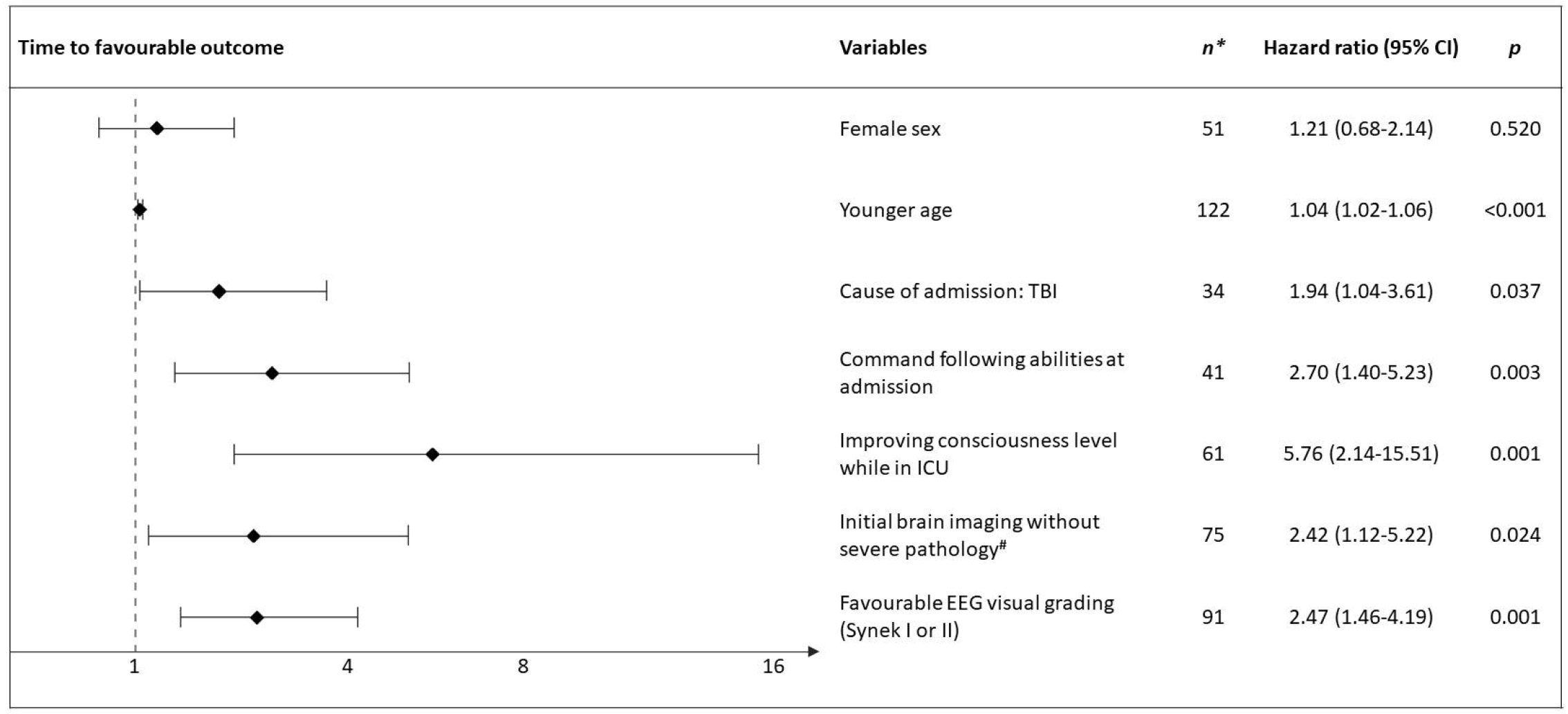
Predictors of time to favourable outcome. This figure depicts independent variables predicting time to favourable outcome (i.e., GOS-E≥4, mRS≤3 and CPC≤2). Death in ICU (*n=*41) was treated as a competing risk in a multivariate Cox proportional regression model. Younger age, patients with TBI, ability to follow commands at admission, improving consciousness level during ICU, no severe pathology at admission brain imaging, and favourable visual grading of EEG (i.e., Synek score I or II) were all independent predictors of earlier recovery. *Of all 123 included patients, one patient without EEG was excluded from this analysis. ^#^Severe pathology on brain imaging was defined as Fisher grade ≥3 (for subarachnoid hemorrhage), Marshall classification ≥3 (for TBI), hemorrhage volume ≥30mL (for intracerebral hemorrhage), strategic hemorrhage or infarct in brainstem (for ischemic stroke or infratentorial hemorrhage), any visible sign of anoxic brain injury on CT scan (for cardiac arrest), global cortical edema (for patients with brain edema), brain tumors with midline compression, compression of basal cisterns and/or signs of hydrocephalus (for patients with any type of brain tumor)

### Machine learning predictive models

Below are results from Random-Forest predictive models, while results from SVM models are presented in the Supplementary file **(Table S1** and **Fig. S1-2)**.

#### EEG-features and functional outcome

Of the predictive models based on individual EEG-features (i.e., Synek score, ABCD categories and P(MCS)), only the Synek score could predict functional outcome at both 3-month (AUC 0.65 [0.61-0.69], PPV 0.43 [0.38-0.47], sensitivity 0.47 [0.44-0.50]) and 12-month (AUC 0.66 [0.63-0.69], PPV 0.64 [0.61-0.68], sensitivity 0.42 [0.42-0.43]). The model based on ABCD categories could not predict 3-month outcome (AUC 0.38 [0.34-0.42], PPV 0.13 [0.08-0.17], sensitivity 0.24 [0.15-0.33]) but could predict 12-month outcome (AUC 0.60 [0.57-0.62], PPV 0.59 [0.50-0.68], sensitivity 0.21 [0.19-0.23]), while the models based on P(MCS) could predict 3-month outcome (AUC 0.64 [0.63-0.67], PPV 0.43 [0.38-0.47], sensitivity 0.47 [0.44-0.50]) but not 12-month outcome (AUC 0.63 [0.61-0.69], PPV 0.32 [0.29-0.34], sensitivity 0.62 [0.58-0.66]) (see also **Table 3** and **Fig. 3**, models I to III). Head-to-head comparison of the same-sample models based on individual EEG-features showed that models based on Synek score outperformed the ABCD model in predicting 3-month outcome (AUC_Synek_ 0.70 [0.69-0.71], AUC_ABCD_ 0.40 [0.33-0.46]) and the P=(MCS) model at 12-month outcome (AUC_Synek_ 0.73 [0.71-0.75], AUC_P(MCS)_ 0.54 [0.50-0.59]) (see also **Table 3** and **Fig. 4**, model Ia compared to model IIa and IIIa).

**Table 3.**
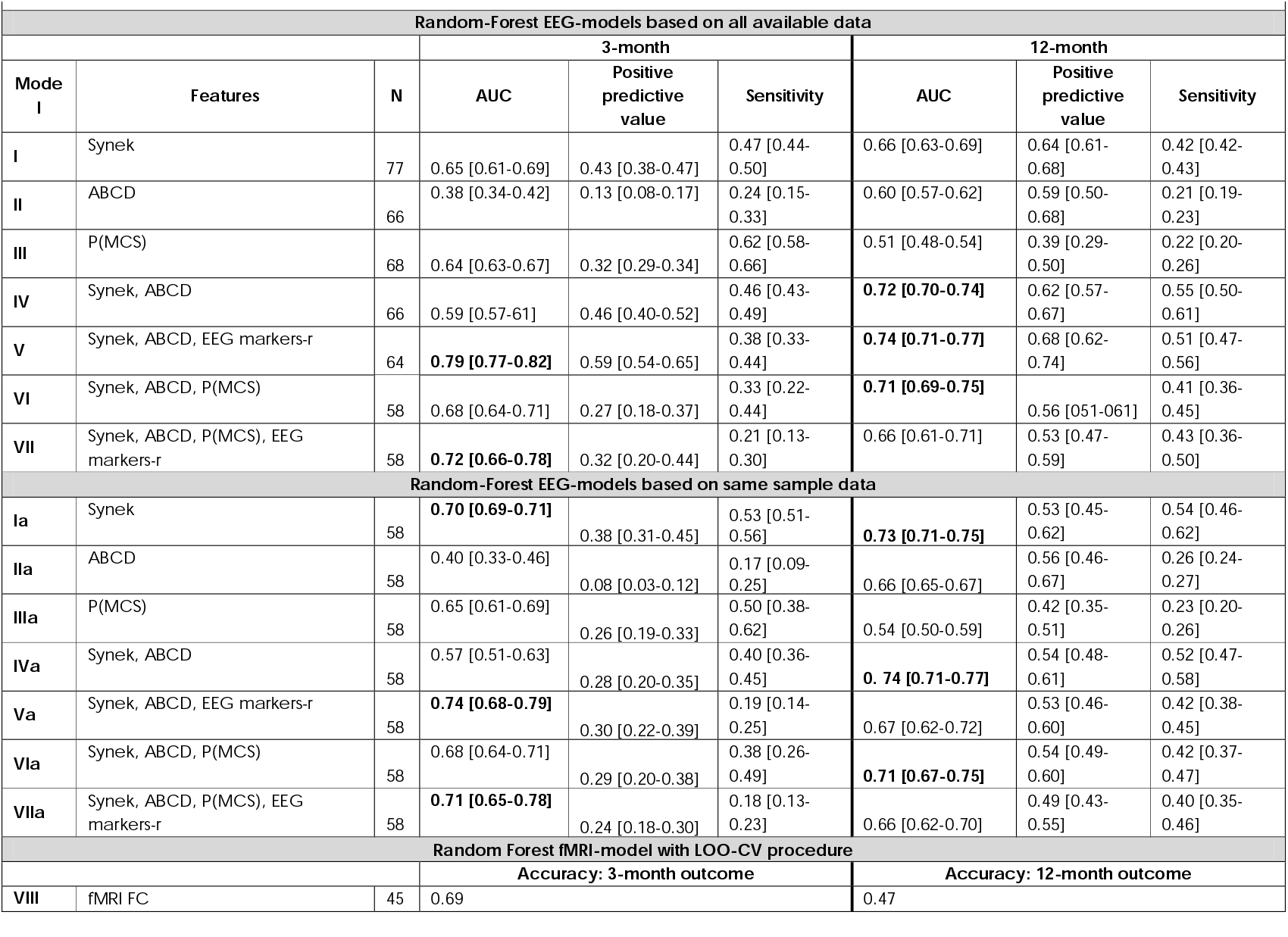
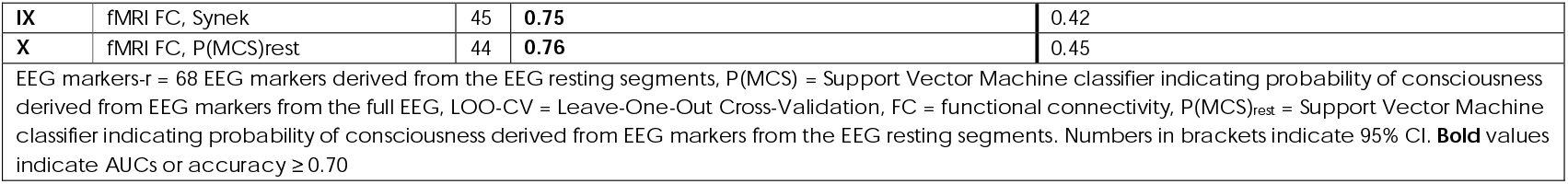
Prediction performance of EEG- and fMRI-features in predicting 3- and 12-month functional outcome.

**Figure 3.**
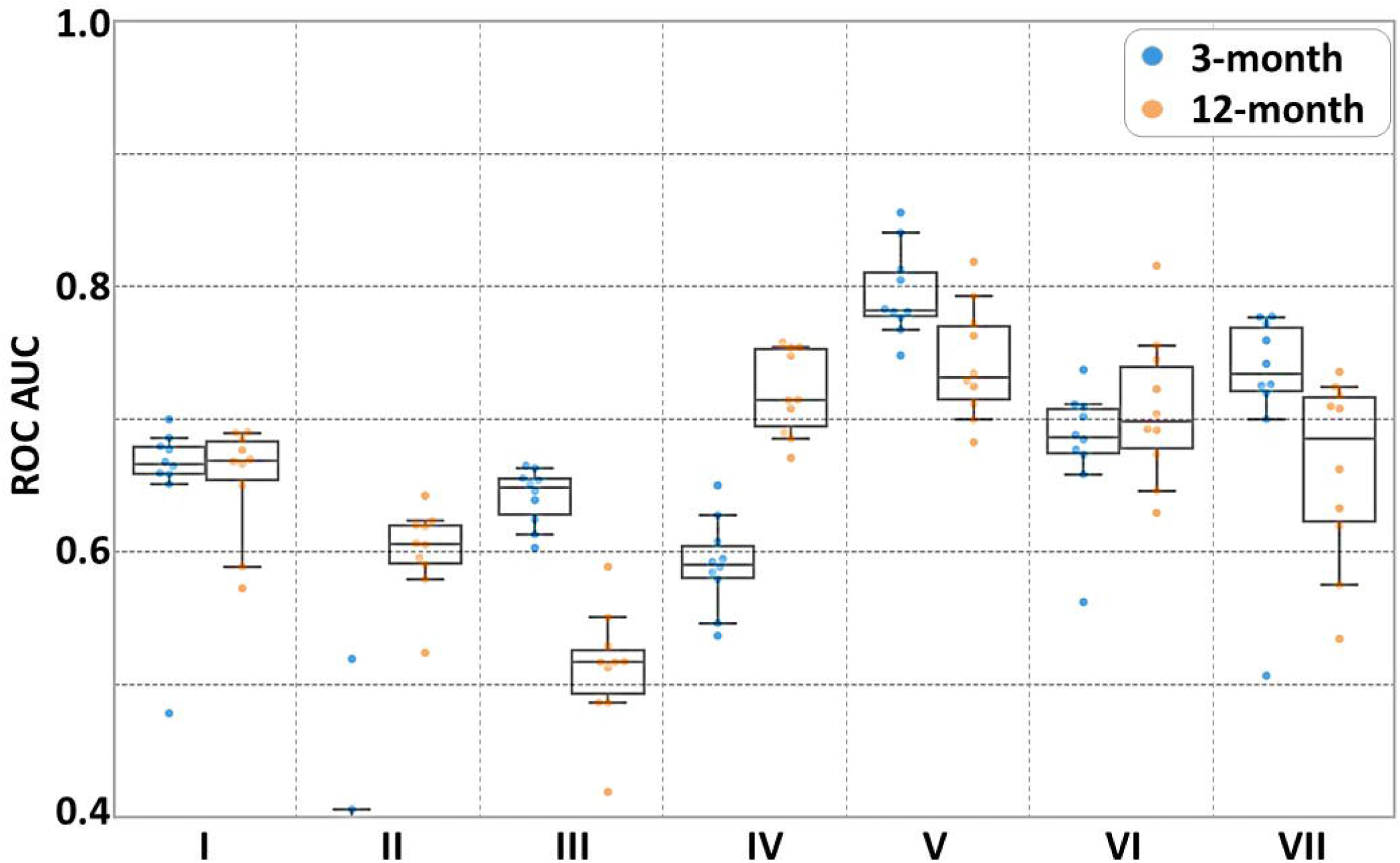
Random-Forest EEG models with maximum available data predicting 3- and 12-month outcomes. Boxplots illustrating model performances (AUCs) of RF-models based on EEG-features predicting 3-month (**blue**) and 12-month (**orange**) functional outcomes. Each model is based on the maximum amount of data available (see also **Fig. 1**). Of the unimodal models (**I-III**), only model **I** based on the Synek score could predict both 3- and 12-month outcomes. The highest AUC for predicting both outcomes (AUC_3-month_ 0.79 [0.77-0.82]; AUC_12-month_ 0.74 [0.71-0.77]) were obtained with the combined model (**V**) based on combination of three EEG-features (i.e., Synek score, ABCD categories and EEG markers-r derived from the SVM consciousness classifier). Overall, this figure shows that while Synek score was the only unimodal EEG-model that predicted both 3- and 12-month functional outcomes, all models based on a combination of EEG-features (**IV-VII**) could predict both 3- and 12-month outcomes with AUCs above chance level. A similar pattern was observed for SVM machine-learning models (see **Fig. S1**). *Individual EEG Random-Forest models:* **I**=Synek, **II**=ABCD, **III**=P(MCS) C. *Combined EEG Random-Forest models*: **IV**=Synek + ABCD, **V**=Synek + ABCD + EEG markers-r, **VI**= Synek + ABCD + P(MCS) and **VII**= Synek + ABCD + P(MCS) + EEG markers-r

**Figure 4.**
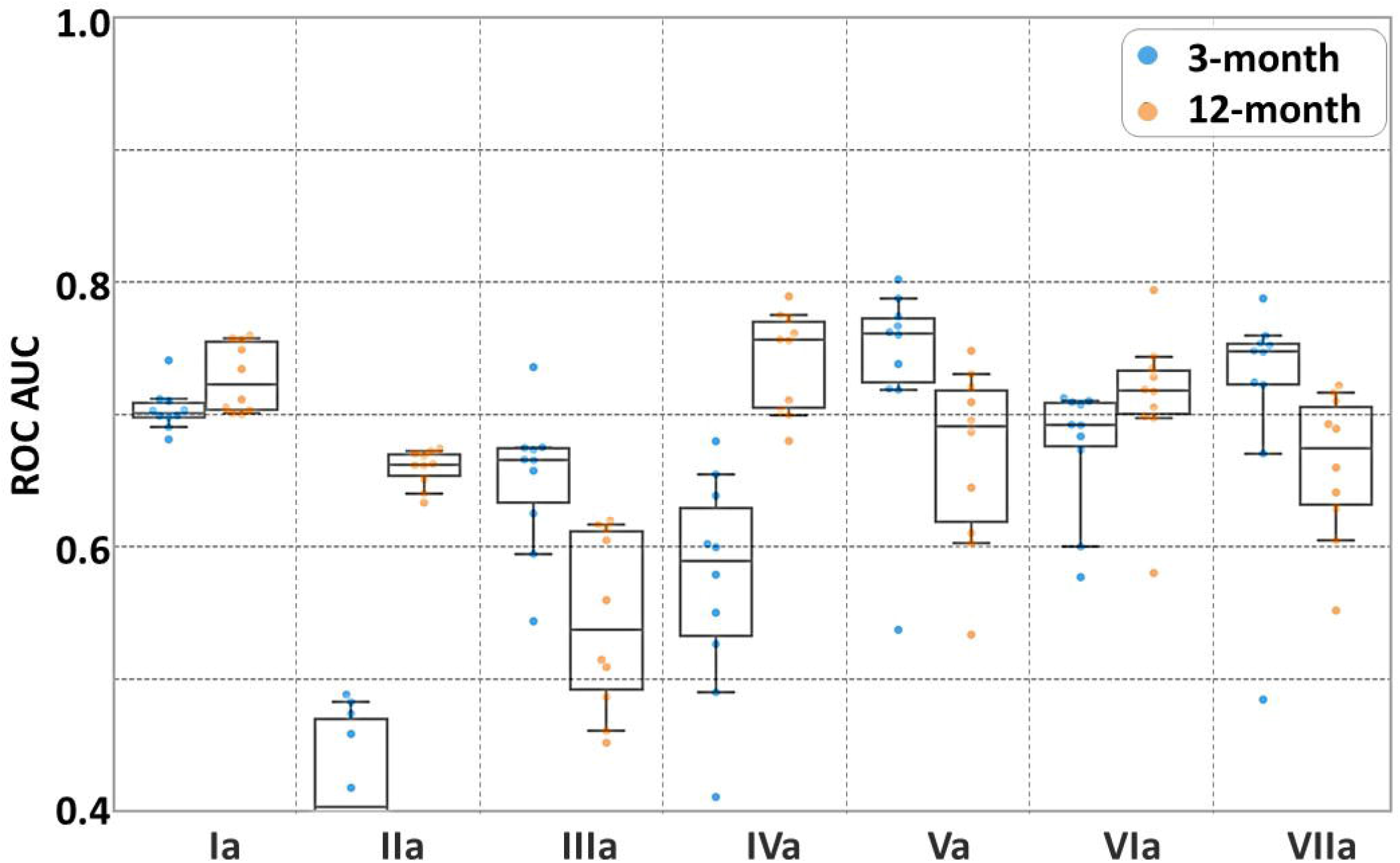
Random-Forest EEG models with same sample data predicting 3- and 12-month outcomes. Boxplots illustrating model performances (AUCs) of machine-learning models based on EEG-features predicting 3-month (**blue**) and 12-month (**orange**) functional outcomes. Each model is based on the same samples (*n=*58) for head-to-head comparison of EEG-features. Of the unimodal models (**Ia-IIIa**), model **Ia** based on Synek score outperformed model **IIa** based on ABCD categories in predicting 3-month outcome (AUC_Synek_ 0.70 [0.69-0.74] vs. AUC_ABCD_ 0.38 [0.31-0.45]). In predicting 12-month outcome, model **Ia** outperformed model **IIIa** which was based on P(MCS) measures (AUC_Synek_ 0.70 [0.69-0.74] vs. AUC_P(MCS)_ 0.54 [0.50-0.59]). Of the combined models based on at least three EEG features (**Va-VIIa**), all models could predict 3- and 12-month outcomes, and none outperformed the others. A similar pattern was observed for SVM machine-learning models (see **Fig. S2**). *Individual same-sample EEG Random-Forest models:* **Ia**=Synek, **IIa**=ABCD, **IIIa**=P(MCS) C. *Combined same-sample EEG Random-Forest models*: **IVa**=Synek + ABCD, **Va**=Synek + ABCD + EEG markers-r, **VIa**= Synek + ABCD + P(MCS) and **VIIa**= Synek + ABCD + P(MCS) + EEG markers-r

All models based on different combination of EEG-features (**Fig. 3** and **Table 3**, models IV-VII and IVa-VIIa) could predict functional outcome at both 3- and 12-month follow-up. The best combination of AUC, PPV and sensitivity for prediction of both outcomes was achieved with the model based on the combination of Synek score, ABCD categories and P(MCS) (3-month outcome: AUC 0.79 [0.77-0.82], PPV 0.59 [0.54-0.65], sensitivity 0.38 [0.33-0.44]; 12-month outcome: AUC 0.74 [0.71-0.77], PPV 0.68 [0.62-0.74], sensitivity 0.51 [0.47-0.56]) (see also **Fig. 3** and **Table 3**, model VI). When comparing the combined EEG same-sample models, all the models performed equally well (**Table 2** and **Fig 3**, models IVa-VIIa).

#### fMRI functional connectivity and functional outcome

Due to limited number of samples with both fMRI data and outcome measures (*n=*45, see also **Fig. 1**), predictive models including fMRI FC was tested with LOO-CV procedure (**Table 3 and Table S1**, models VIII-X). fMRI FC measures tested with both random-forest and SVM algorithm showed evidence suggesting that predicting 3-month outcome is possible (random-forest model VIII: accuracy 0.69; SVM model VIII: accuracy 0.78), but not 12-month outcome (random-forest model VIII: accuracy 0.47; SVM model VIII: accuracy 0.47). More samples are required to confirm and correctly estimate the performance of such models.

#### Combined EEG- and fMRI-features and functional outcome

We evaluated prediction of 3- and 12-month functional outcomes with the combination of fMRI FC with Synek score (*n=*45) or P(MCS) derived from EEG markers-r (*n=*44) as depicted in **Table 3** (models IX and X). Both combined models showed evidence that predicting 3-month outcome is possible, with accuracies between 0.73-0.84 but not 12-month outcome (**Table 3 and S1**, models IX and X), regardless of which algorithm was used.

## Discussion

In this, to our knowledge, first prospective multimodal cohort study including 123 ICU patients with acute DoC from various underlying conditions, we show that machine learning algorithms applied to EEG- and fMRI-features obtained soon after ICU admission can predict 3-month functional outcome, while 12-month outcome can only be predicted by EEG-features. Furthermore, we have identified important, readily available, independent predictive clinical variables of time to favourable recovery.

EEG-features in combination, as well as EEG Synek score as an individual model, predicted both 3- and 12-month functional outcomes (**Fig. 3** and **Table 3**), whereas all models based on fMRI functional connectivity measures could only predict 3-month outcome (**Table 3**). EEG recordings were available from 77 patients with outcome measures at both 3 and 12 months, while we only had fMRI sequences from 45 of these patients, thus resulting in a substantially reduced amount of data available for the fMRI-feature models. While the quality of the data that underly machine learning models is crucial, data quantity is also important because datasets with many variables but limited number of samples introduce high level of variance, rendering the models imbalanced.^42^ Despite our relatively large population of acute DoC patients, our results, especially those including fMRI-features, should therefore be interpreted with caution, until further validation from ongoing multicentre studies.^43^ These factors may also explain the relatively low PPV and sensitivities despite high AUCs of the combined EEG models, which were based on data from patients with a complete dataset including all EEG-features (*n=*58).

Despite the abovementioned limitations, we could show that most EEG-features predicted both early and late functional outcomes individually and in various combinations (**Fig. 3** and **Table 3**). This is an important finding because EEG is much more available in the ICU than advanced neuroimaging such as fMRI, and EEG-features like ours can be easily implemented in an ICU setting. When comparing the individual EEG-features head-to-head with the same-sample models (**Fig. 4** and **Table 3**), we found that the Synek score outperformed the ABCD categories for the prediction of short-term outcome and that the SVM classifier derived P(MCS) outperformed the ABCD categories for the prediction of long-term outcome. This finding may be explained by the fact that the Synek score was assessed manually by two board-certified electroencephalographers with many years of experience with ICU EEG, while the ABCD and P(MCS) features were developed in patient groups that differed from ours (i.e., homogenous cardiac arrest^35^ and chronic DoC cohorts^36^ vs. acute DoC cohort with heterogeneous brain injuries). Furthermore, visual analysis of EEGs is routinely used for prognostication in ICU populations like the present cohort, which may also explain the higher performance of the models based on Synek score. Still, we could show that combining different EEG-features resulted in the best predictive performance of the models, regardless of the algorithm used (**Table 3** and **Table S1**). These are important findings because most ICU sites with acute DoC patients do not have the resources to perform advanced EEG assessment using machine learning classifiers. These sites can thus safely rely on experienced electroencephalographers using established criteria for visual EEG analyses instead. If the necessary electroencephalographer expertise is unavailable, however, external data-driven analysis of EEGs may become a suitable option for those sites in the near future.

Models including fMRI-features were tested with a LOO-CV procedure due to the limited number of available samples. Results indicate that fMRI FC both alone and in combination with some EEG-features may be useful to predict early functional outcome at 3 months (**Table 3**), but not (yet) late outcome at 12 months. The LOO-CV procedure limits data waste and is therefore primarily used for small datasets, but a major limitation is that the results are prone to optimistic interpretation and therefore need external validation in larger datasets.^40^

In the first paper from the CONNECT-ME study,^18^ we found that EEG and fMRI-features predicted levels of consciousness of acute DoC patients at the time of ICU discharge. Importantly, EEG and fMRI were performed without active consciousness paradigms, thus patients likely had different degrees of residual consciousness (e.g., including those who could not have participated in active paradigms^44^). Collectively, our findings indicate that both EEG and fMRI have the potential not only to predict level of consciousness during ICU admission^18^ but also to predict functional outcome of patients with brain injury of various causes resulting in acute DoC in the early phase of hospitalization and (EEG, at least) up to one year after discharge from the ICU.

In line with a recent study about recovery trajectories of patients with cognitive motor dissociation,^14^ we additionally identified readily available clinical features as independent predictors of time to favourable functional outcome (**Fig. 2**). In our heterogenous patient cohort reflecting a real-life ICU setting, we confirmed that TBI is related to earlier recovery. Furthermore, patients who were younger, could follow commands at ICU admission, had no severe pathology on initial brain imaging, and showed improving consciousness level in the ICU, also recovered earlier. Similarly, patients with favourable functional outcomes at 3 and 12 months were more likely to be discharged directly to their own home, while patients with unfavourable outcome were more often discharged to rehabilitation facilities and nursing homes (**Table 2**). This is explained by the fact that patients with more severe injuries needed higher level of care and were thus discharged to facilities with higher level of rehabilitation resources. All these findings can help clinicians when guiding patient families about the prospects of recovery, including the time it takes to achieve a good recovery.

### Strengths and limitations

Several limitations need to be considered. As a single center study, CONNECT-ME is susceptible to sampling bias. A relatively large number of patients (33%) died in the ICU, most due to withdrawal of life-sustaining therapy because of a presumed poor prognosis. While the current study included 123 patients, data from only 77 patients were available for the final analysis of 12-month outcomes. Thus, the remaining cohort with available follow-up data consisted of patients who were expected to regain better functional outcome. This skewed the dataset used in the machine learning models. The predictive performance of these models may hence have been biased in that they lacked the (potential) clinical trajectories of patients who had life-sustaining therapy withdrawn. To account for this bias to some extent, in our analysis of independent variables related to time to favourable outcome we included in-hospital death as a competing risk in the multivariate Cox proportional hazards regression model. Still, death due to withdrawal of life-sustaining therapy in the ICU remains an important limitation and cannot be fully accounted for when studying ICU-patients with acute severe brain injury and DoC. Since EEG is more available in ICU than fMRI, it is routinely used for prognostication of acute DoC patients, especially of those admitted post-cardiac arrest.^45^ Excluding patients who died in the ICU may therefore have decreased the performance of the EEG models as well.

MRI scans are logistically very challenging to obtain in the ICU and are thus less often performed than EEG, which might be yet another selection bias, affecting the fMRI models owing to exclusion of patients without available fMRI. However, in our cohort, we found no statistically significant difference in the frequency with which fMRI was performed when comparing patients who died in the ICU to those who were discharged alive (**Table 1**), or when comparing patients with favourable outcome to those with unfavourable outcome (**Table 2**), suggesting this might be of lesser importance to the overall results. Our study population is a heterogeneous group of patients with various causes of DoC, rendering subgroup analysis unreliable due to the low number of patients in each group. Thus, further validation is needed to confirm our findings.

On the positive side, our findings are generalizable to a real-life ICU setting and acute DoC patients with various causes of brain injury. We also evaluated functional outcome in our cohort by using three different outcome scales designed for stroke (mRS),^37^ TBI (GOS-E)^46^ and cardiac arrest (CPC)^39^ patients to account for the heterogenicity of our patients. Owing to logistical challenges and resources needed for advanced data analyses, to our knowledge, no previous EEG/fMRI study has managed to investigate acute DoC in a larger ICU cohort or with a longer follow-up than ours.

## Conclusion

We show that EEG early during ICU admission predicted both 3- and 12-month functional outcomes of acute DoC patients with various causes of brain injury, and that fMRI resting-state measures might be useful to predict 3-month outcome. Furthermore, young age, TBI, initial brain imaging without severe pathology, ability to follow commands during ICU admission, improving consciousness level during the ICU stay, as well as favourable visual EEG grading, all independently predicted shorter time to favourable functional outcome. In sum, we suggest that combining EEG- and fMRI-based machine learning models with readily available clinical data allows for reliable short-term outcome prediction of patients with coma and other acute DoC and potentially can predict long-term outcome up to one year after ICU discharge.

## Abbreviations

AN: auditory network
AUC: area under the curve
BOLD: blood oxygen level dependent
CONNECT-ME: consciousness in neurocritical care cohort study using fMRI and EEG
CPC: cerebral performance category
CS: confusional state
DMN: default mode network
DoC: disorders of consciousness
eMCS: emerging from MCS
FC: functional connectivity
fMRI: functional MRI
FOUR: full outline of unresponsiveness
FPN: frontoparietal network
GCS: Glasgow coma scale
GOS-E: Glasgow outcome scale-extended
ICU: intensive care unit
MCS: minimally conscious state
mRS: modified Rankin scale
P(MCS): probability of being at least minimally conscious derived by the SVM classifier from the full EEG recording
SMN: sensorimotor network
SN: salience network
SVM: Support Vector Machine
TBI: traumatic brain injury
UWS: unresponsive wakefulness syndrome
VN: visual network
WLST: withdrawal of life-sustaining therapy

## Acknowledgements

We are very grateful to the patients and their families who participated in this study. Additionally, we thank the staff at the departments of clinical neurophysiology, radiology, cardiology, neurology, and intensive care units (sections 6062, 2143, 4131 and 4141), at Rigshospitalet, Copenhagen University Hospital, for their cooperation and support. A special thanks to neurointensive care nurse Ida Møller for her help with screening patients, and to neurophysiology technicians Puk Overgaard Christiansen, Rikke Dresner, Julie Dyppel, Dorthe Husum Hansen, Allan Rene Hansen, Lars Hvidtfeldt, Sundus Said Muse Issa, Trine Jessen, Camilla Hedegaard Mundt Larsen, Pia Christina Stade-Larsen, Marianne Sonnichsen, and Parisa Yazdanyar for their help with EEG recordings. Figure 1 were created with Biorender.com.

## Funding

Funding was supplied by Region Hovedstadens Forskningsfond, Lundbeck Foundation, Rigshospitalets Forskningspuljer and Offerfonden. Material execution, content and results are the sole responsibility of the authors. The assessments and views expressed in the material are the authors’ own and are not necessarily shared by the funders.

## Notes

### Competing Interest Statement

The authors have declared no competing interest.

### Clinical Protocols

https://www.ncbi.nlm.nih.gov/pmc/articles/PMC6278242/

https://academic.oup.com/brain/article/146/1/50/6696511?login=false

