## Supplemental files for "Multimodal prediction of 3- and 12-month outcomes in ICU-patients with acute disorders of consciousness"

**Supplementary data**

1. **Box S1:** Functional outcome scales
2. **Table S1:** Prediction performance of SVM models predicting 3- and 12-month functional follow-up
3. **Figure S1:** Boxplots of SVM unimodal and multimodal models based on maximum available data
4. **Figure S2:** Boxplots of SVM same-sample models based on full-set data

| **Box S1. Functional outcome scales** | | |
| --- | --- | --- |
| **Scale** | **Categories/levels** | **Explanation** |
| **Modified Rankin scale (mRS)** | 0: No current symptoms | - |
|  | 1: No significant disability | Able to carry out all usual activities despite minor symptoms |
|  | 2: Slight disability | Unable to carry out all usual activities, but able to look after daily affairs without help from others |
|  | 3: Moderate disability | Require some help but can walk unassisted |
|  | 4: Moderately severe disability | Assistance needed to attend bodily needs. Unable to walk unassisted |
|  | 5: Severe disability | Constant care needed. Patient is bedridden |
|  | 6: Dead | - |
| **Glasgow Outcome Scale Extended**  **(GOS-E)** | 1: Dead | - |
|  | 2: Persistent vegetative state | Only reflex behavior, inability to communicate |
|  | 3: Severe disability (lower) | Requires frequent help from other most of the time daily |
|  | 4: Severe disability (upper) | Can take of oneself alone for minimum 8 hours per day |
|  | 5: Moderate disability (lower) | Able to work in sheltered workshop or not at all |
|  | 6: Moderate disability (upper) | Work capacity reduced. Social activities reduced to less than 50% of preinjury status |
|  | 7: Good recovery (lower) | Minor issues effecting daily living |
|  | 8: Good recovery (upper) | No current problems effecting daily living |
| **Cerebral Performance Category (CPC)** | 1: Good cerebral performance | Conscious, alert and able to work despite minor deficits. |
|  | 2: Moderate cerebral disability | Conscious, independent of other for daily life activities. Can work in sheltered environment |
|  | 3: Severe cerebral disability | Conscious, dependent on other for daily support. Can range from ambulatory to severe dementia/paralysis |
|  | 4: Coma or vegetative state | Unaware without any ability to interact with the environment |
|  | 5: Dead/Brain dead | - |
| Components of the different functional outcome scales. The green lines show threshold between favorable and unfavorable outcome for each scale | | |

| **Table S1** Prediction performance of EEG and fMRI-features in predicting 3- and 12-month functional outcome | | | | | | | | |
| --- | --- | --- | --- | --- | --- | --- | --- | --- |
| **Support Vector Machine EEG-models based on all available data** | | | | | | | | |
|  | | | **3-month** | | | **12-month** | | |
| **Model** | **Features** | **N** | **AUC** | **Positive predictive value** | **Sensitivity** | **AUC** | **Positive predictive value** | **Sensitivity** |
| **I** | Synek | 77 | 0.68 [0.67-0.70] | 0.17 [0.12-0.23] | 0.38 [0.26-0.50] | 0.59 [0.57-0.62] | 0.13 [0.09-0.17] | 0.26 [0.17-0.34] |
| **II** | ABCD | 66 | 0.39 [0.37-0.42] | 0.12 [0.08-0.17] | 0.41 [0.27-0.55] | 0.62 [0.61-0.63] | 0.65 [0.57-0.73] | 0.22 [0.21-0.22] |
| **III** | P(MCS) | 68 | 0.67 [0.66-0.68] | 0.30 [0.26-0.34] | 0.68 [0.59-0.76] | 0.56 [0.53-0.59] | 0.35 [0.30-0.40] | 0.54 [0.44-0.64] |
| **IV** | Synek, ABCD | 66 | 0.63 [0.60-0.66] | 0.23 [0.19-0.28] | 0.42 [0.32-0.52] | 0.61 [0.57-0.66] | 0.53 [0.48-0.59] | 0.34 [0.29-0.39] |
| **V** | Synek, ABCD, EEG markers-r | 64 | **0.71 [0.67-0.75]** | 0.42 [0.36-0.49] | 0.50 [0.46-0.55] | **0.71 [0.67-0.74]** | 0.55 [0.54-0.57] | 0.56 [0.52-0.60] |
| **VI** | Synek, ABCD, P(MCS) | 58 | **0.70 [0.66-073]** | 0.14 [0.09-0.18] | 0.20 [0.15-0.24] | 0.68 [0.64-0.71] | 0.39 [0.32-0.46] | 0.36 [0.28-0.43] |
| **VII** | Synek, ABCD, P(MCS), EEG markers-r | 58 | 0.66 [0.62-0.70] | 0.41 [0.34-0.48] | 0.47 [0.40-0.54] | **0.70 [0.65-0.76]** | 0.37 [0.28-0.46] | 0.51 [0.41-0.6.] |
| **Support Vector Machine same sample EEG-models** | | | | | | | | |
| **Ia** | Synek | 58 | **0.70 [0.69-0.71]** | 0.07 [0.03-0.11] | 0.11 [0.06-0.16] | 0.47 [0.39-0.55] | 0.30 [0.22-0.39] | 0.56 [0.46-0.67] |
| **IIa** | ABCD | 58 | 0.43 [0.39-0.48] | 0.00 [0.00-0.01] | 0.02 [-0.03-0.07] | 0.67 [0.66-0.68] | 0.60 [0.51-0.68] | 0.31 [0.27-0.34] |
| **IIIa** | P(MCS) | 58 | **0.71 [0.70-0.73]** | 0.08 [0.03-0.13] | 0.14 [0.08-0.20] | 0.63 [0.59-0.68] | 0.38 [0.30-0.46] | 0.57 [0.47-0.66] |
| **IVa** | Synek, ABCD | 58 | 0.64 [0.61-0.67] | 0.05 [0.01-0.10] | 0.12 [0.02-0.22] | 0.65 [0.61-0.69] | 0.46 [0.35-0.57] | 0.34 [0.28-0.40] |
| **Va** | Synek, ABCD, EEG markers-r | 58 | **0.70 [0.64-0.76]** | 0.32 [0.24-0.39] | 0.43 [0.36-0.50] | 0.65 [0.61-0.69] | 0.41 [0.34-0.49] | 0.46 [0.41-0.52] |
| **VIa** | Synek, ABCD, P(MCS) | 58 | **0.70 [0.66-0.73]** | 0.14 [0.09-0.18] | 0.20 [0.15-0.24] | 0.67 [0.64-0.71] | 0.39 [0.32-0.46] | 0.36 [0.28-0.42] |
| **VIIa** | Synek, ABCD, P(MCS), EEG markers-r | 58 | **0.70 [0.65-0.76]** | 0.37 [0.28-0.46] | 0.51 [0.41-0.61] | 0.66 [0.62-0.70] | 0.41 [0.34-0.48] | 0.47 [0.40-0.54] |
| **Support Vector Machine fMRI-model with LOO-CV procedure** | | | | | | | | |
|  |  |  | **Accuracy: 3-month outcome** | | | **Accuracy: 12-month outcome** | | |
| **VIII** | fMRI FC | 45 | **0.78** | | | 0.47 | | |
| **IX** | fMRI FC, Synek | 45 | **0.73** | | | 0.42 | | |
| **X** | fMRI FC, P(MCS)rest | 44 | **0.84** | | | 0.61 | | |
| EEG markers-r = 68 EEG markers derived from the EEG resting segments, P(MCS) = Support Vector Machine classifier indicating probability of consciousness derived from EEG markers from the full EEG, LOO-CV = Leave-One-Out Cross-Validation, FC = functional connectivity, P(MCS)_rest_ = Support Vector Machine classifier indicating probability of consciousness derived from EEG markers from the EEG resting segments. Numbers in brackets indicate 95% CI. **Bold** values indicate AUCs or accuracy ≥ 0.70 | | | | | | | | |

**Figure S1. Support Vector Machine EEG models with maximum available data predicting 3- and 12-month outcomes.** Boxplots illustrating model performances (AUCs) of SVM-models based on EEG-features predicting 3-month (**blue**) and 12-month (**orange**) functional outcomes. Each model is based on the maximum amount of data available (see also **Fig. 1**). Of the unimodal models (**I-III**), both model **I** and **III** based on the Synek score and P(MCS), respectively, could predict both 3- and 12-month outcome. Of the combined models (Models **IV-VII**) the highest AUC for predicting both outcomes were obtained with models **V-VII** based on all three EEG-features (i.e., Synek score, ABCD categories, P(MCS) and/or EEG markers-r). In sum, this figure shows that while not all unimodal EEG models could predict both 3- and 12-month functional outcomes, all models based on a combination of EEG-features (**IV-VII**) could predict both 3- and 12-month outcomes with AUCs above chance level. *Individual EEG SVM-models:* **I**=Synek, **II**=ABCD, **III**=P(MCS) C. *Combined EEG SVM-models*: **IV**=Synek + ABCD, **V**=Synek + ABCD + EEG markers-r, **VI**= Synek + ABCD + P(MCS) and **VII**= Synek + ABCD + P(MCS) + EEG markers-r

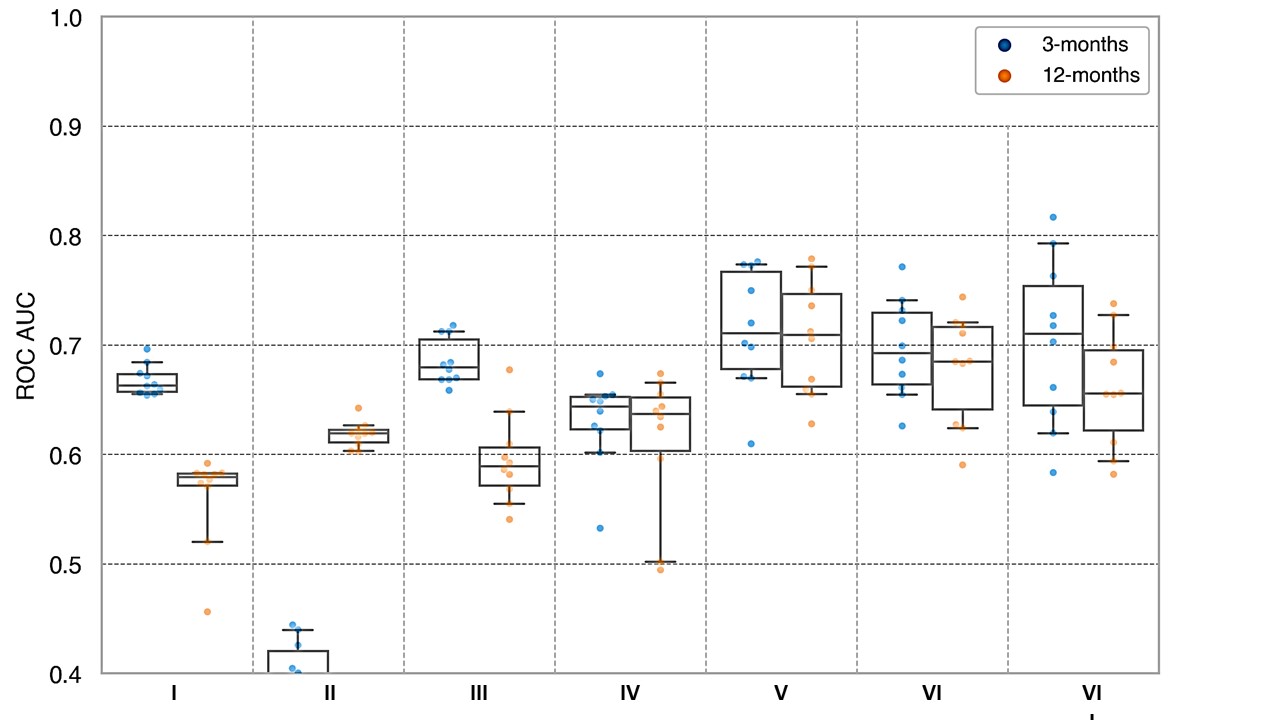

**Figure S2. Support Vector Machine EEG models with same sample data predicting 3- and 12-month outcomes.** Boxplots illustrating model performances (AUCs) of machine-learning models based on EEG-features predicting 3-month (**blue**) and 12-month (**orange**) functional outcomes. Each model is based on the same samples (n=58) for head-to-head comparison of EEG-features. Of the unimodal models (**Ia-IIIa**), model **Ia** (i.e., Synek model) and **IIIa** (i.e., P(MCS) model) outperformed model **IIa** (i.e., ABCD model) in predicting 3-month outcome (AUC_Synek_ 0.70 [0.69-0.71] vs. AUC_P(MCS)_ 0.71 [0.70-0.73] vs. AUC_ABCD_ 0.43 [0.39-0.48]). In predicting 12-month outcome, model **IIa** (i.e., ABCD model) and **IIIa** outperformed model **Ia** (Synek model) (AUC_P(MCS)_ 0.63 [0.59-0.68] vs. AUC_ABCD_ 0.67 [0.66-0.68] vs. AUC_Synek_ 0.47 [0.39-0.55]). Of the combined models (**Va-VIIa**), all models could predict 3- and 12-month outcomes, and none outperformed the others. *Individual same-sample EEG SVM-models:* **Ia**=Synek, **IIa**=ABCD, **IIIa**=P(MCS) C. *Combined same-sample EEG SVM-models*: **IVa**=Synek + ABCD, **Va**=Synek + ABCD + EEG markers-r, **VIa**= Synek + ABCD + P(MCS) and **VIIa**= Synek + ABCD + P(MCS) + EEG markers-r

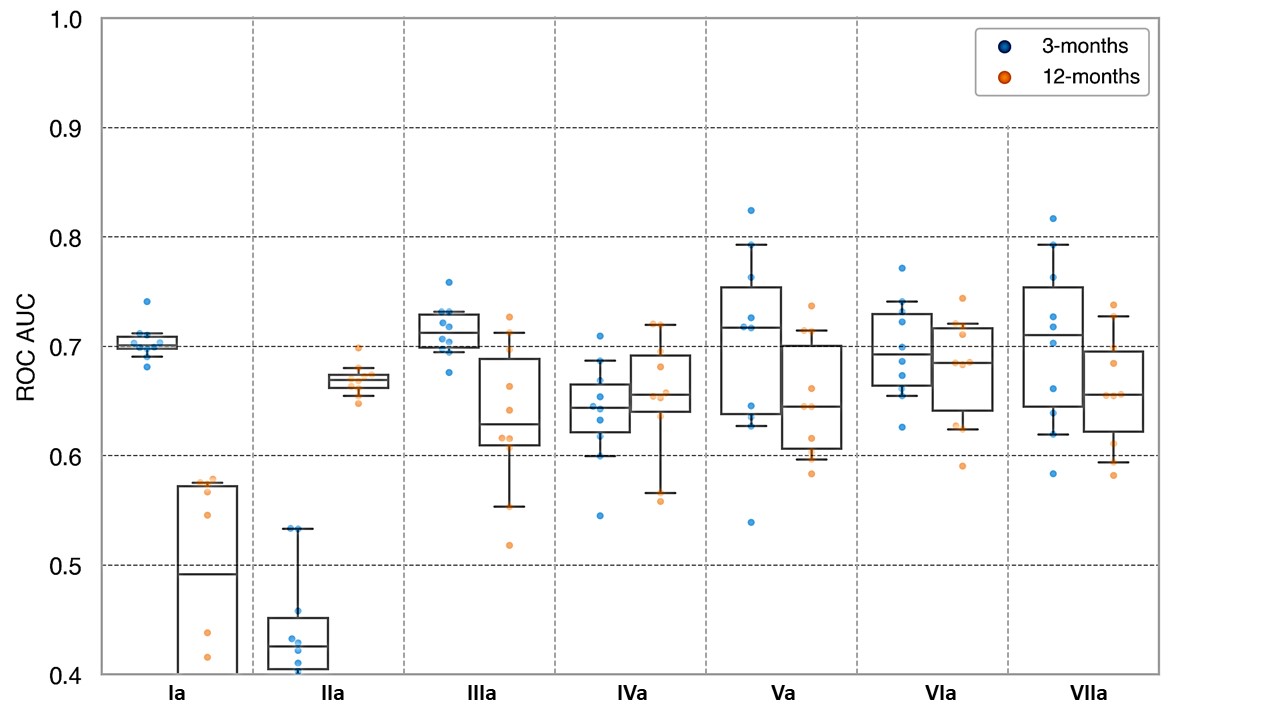
